# Spatiotemporal Correlations of Mesoscale Neocortical Activities in Patients with Focal Epilepsy

**DOI:** 10.1101/2021.03.31.21254715

**Authors:** Somin Lee, Sarita S. Deshpande, Edward M. Merricks, Michel J.A.M. van Putten, Catherine A. Schevon, Wim van Drongelen

**Author notes:** Corresponding Authors Wim van Drongelen and Catherine A. Schevon, or. S.L, S.S.D., C.A.S., and W.v.D. contributed equally to this work. **Author Contributions** Author contributions: S.L, S.S.D., W.v.D. designed research. S.L., S.S.D., M.J.A.M.v.P., C.A.S., E.M.M., W.v.D. performed research. S.L., S.S.D., E.M.M., M.J.A.M.v.P., W.v.D. analyzed data. S.L., S.S.D., M.J.A.M.v.P., C.A.S., W.v.D. wrote manuscript.

## Abstract

Brain function is reflected in both the action potentials of individual neurons and interactions through e.g. synaptic currents reflected in widespread, slow fluctuations of the local field potential (LFP). We analyzed microelectrode array data to determine state-dependent correlations between action potential and LFP during seizure events as well as interictally in patients with focal epilepsy. We also examined activity in two different cortical network territories: the seizure core and surrounding penumbra (Schevon et al., 2012).

The cross-correlation of spiking activity in the core showed an association of the ictal action potentials with the global oscillatory aspect of the seizure activity and indicated local failure of inhibitory restraint surrounding the ictal spike. These patterns were not observed in the penumbra.

Our analyses from clinical recordings and a model of a single ictal spike in the core revealed that both the temporal and spatial components of the network’s cross-correlation can be approximated by a sine cardinal (*sinc*) function. The biological interpretation of these findings is that important functional differences across the neocortical network exist, with a critical role of the millimeter-range excitatory connections within the grey matter. Therefore, localized intervention that prevents escape of hyperactivity from the seizure core may be considered as a therapeutic strategy.

**Significance Statement:** Analysis and modeling of spatiotemporal cross-correlation between multi-unit spike activity and the local field potential of human seizure activity demonstrates a difference of network functionality between the seizure core and its surrounding penumbral network. In this study, we show that millimeter-range excitatory connections, known to exist within the gray matter of neocortex, play a critical role in propagating the effects of ictal hyperexcitation. These findings contribute to our understanding of ictal mechanisms and hold promise for further development of strategies to detect the ictal core territory and prevent seizure propagation.

## Introduction

Spatiotemporal patterns of brain electrical activity are used to study mechanisms underpinning brain activity and are often employed to guide therapeutic approaches in neurology. During surgical evaluation of patients with epilepsy, brain electrical activity may be obtained at different scales: global activity recorded by macroelectrodes at the scalp or directly on the cortex as well as meso- and microscale activities using arrays or bundles of microelectrodes (1–3). Although the currently available data set of recordings in patients and animal epilepsy models have significantly increased our knowledge of the interictal and ictal processes, the details of the spatiotemporal patterns that play role in neural activity across cortical networks are not well understood.

One critically important question in understanding cortical dynamics is how activities of individual neurons relate to local and overall network function observed in between and during seizures. Recent studies (2–4) have quantified the interaction of neural networks during human focal seizures across micro-, meso- and macroscopic scales. One study showed that the spike-triggered average (STA) of the ongoing low frequency component of the local field potential (*LFP*), i.e., the cross-correlation between action potential and its surrounding compound activity, was characterized by a function strongly resembling a sine cardinal (*sinc*) function (4). The Fourier transform of the *sinc* function is a rectangular function in the frequency domain (5). Accordingly, the measured ictal train of action potentials passed through a filter with a rectangular (ideal) frequency characteristic generated an output that correlates well with the observed seizure (4). While the ictal STA was determined in the time and frequency domains, the spatial aspect of the details of the correlation between action potential and ongoing low frequency component of the neural activity remains unknown. Understanding this relationship is especially of interest since the overall low frequency component of the field potential is captured and used in the clinical EEG recordings of the seizure activity. Consequently, a better understanding of its relationship to underlying neuronal firing might significantly improve insight into the seizure mechanisms, as well as diagnostics and interpretability of EEG recordings in clinical settings.

The purpose of this work is to extend our knowledge base by investigating the ictal and interictal cross-correlations between action potential activity and the ongoing low frequency component of the neural activity and by adding a spatial component to these correlations. We achieve this by analyzing data obtained from microelectrode array recordings during human focal seizures across different cortical territories combined with a theoretical-modeling approach. We evaluate the outcome of our analysis in the context of network properties across neocortical territories and we discuss the relationship between these spatial properties and their potential significance for therapeutic approaches.

## Results

Human cortical activity before, during, and after focal seizures was analyzed using recordings obtained from 96-channel, 4×4mm micro-electrode arrays (MEA). We determined local multi-unit neural firing activity and the associated low frequency component of the local field potential (*LFP*) of the surrounding network (e.g., Movie S1). We describe the temporal and spatiotemporal effects of the neuronal action potential (spike) on the surrounding cortical network during the interictal state (>2 hours away from the seizure) as well as the ictal phase. We consider a cortical network located in both the core territory and its surrounding penumbra, territories which have been described previously (1).

We first introduce a model of an electrode that measures the network response to a single spike. Next, we describe the relationship between spike events and the *LFP* by presenting temporal cross-correlations/spike-triggered averages (STAs). Finally, we describe the spatiotemporal cross-correlation in both cartesian and polar coordinates (Fig. S1, Methods).

### Model of macroelectrode recording predicts sinc function in spatial domain

To generate a theoretical prediction of the spatial pattern of the LFP associated with spiking activity, we extended the findings (4) by analyzing a spatiotemporal model of ictal activity as seen by a macroelectrode (Fig. 1). In this model, a single ictal action potential is generated at the center under a macroelectrode that covers a cortical surface, and the associated field potential is measured. If the spike is represented by a delta function, *δ*(*r, τ*), and the ictal network under the electrode is modeled as a linear time-invariant (LTI) system, this setup allows measurement of the correlation between spike and *LFP* as the system’s unit impulse response, *UIR*(*r, τ*). The *UIR* measured by the electrode is governed by the action potential’s associated ictal network activity represented by some unknown function *f*(*r, τ*) of space (*r*) and time (*τ*).

We first evaluate the network’s unit impulse response in the time domain. Because the potential of cortical generators attenuates sharply with distance, we assume that we may ignore contributions from activity in areas not directly under the macroscopic electrode. Under this assumption, the electrode’s signal can be approximated by summing the contributions over the neocortical area under the electrode:

**Figure 1:**
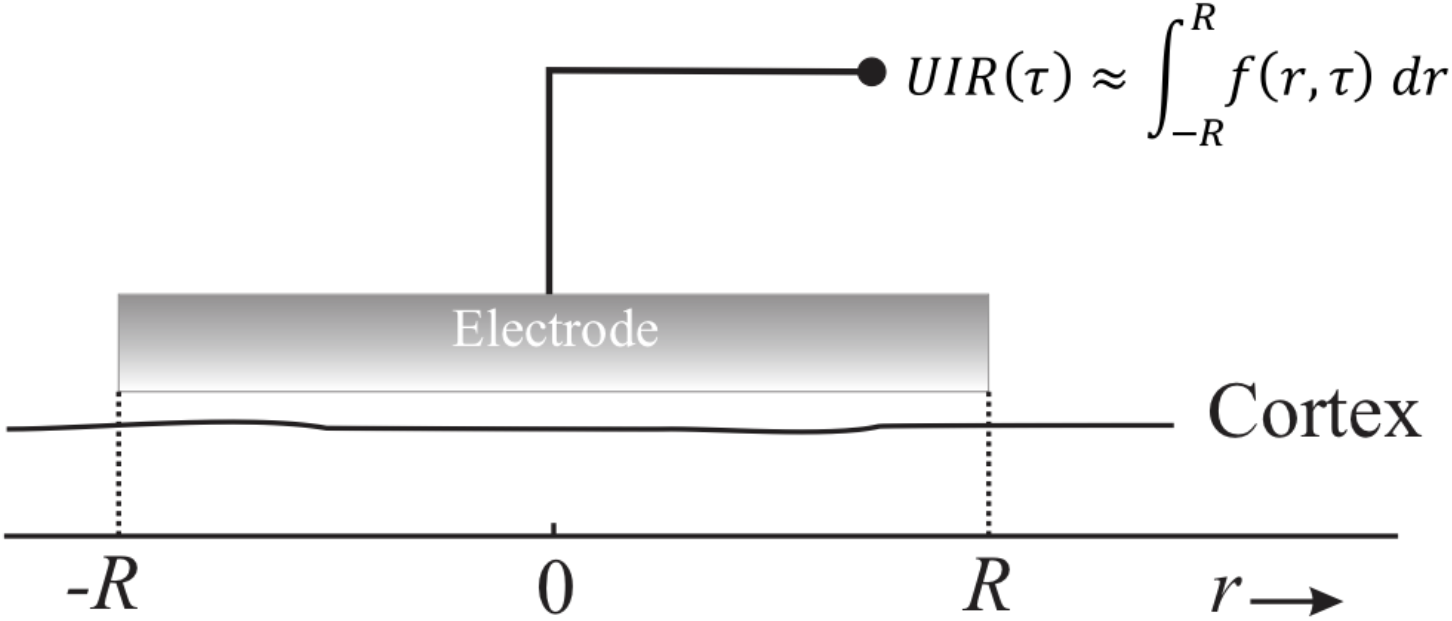
A model of a network’s unit impulse response measured by a macroelectrode. The electrode covers an area of one-dimensional cortex where we record the effect of a single centrally located ictal action potential, *δ*(*r, τ*) = *δ*(0,0), i.e., the macroelectrode measures the underlying network’s temporal unit impulse response *UIR*(*τ*). This measurement can be approximated by an unknown action potential’s spatiotemporal cortical activation function *f*(*r, τ*), integrated over the domain [−*R, R*] covered by the electrode.

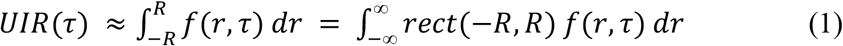

Using results from Eissa et al., (2018), we may substitute *UIR*(*τ*)with the following relationship:

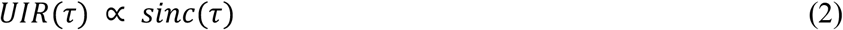

Because the *sinc* function is defined as the Fourier transform of a rectangular function, the relationship between time and space parallels a time-frequency Fourier-transform-pair.

Accordingly, we find:

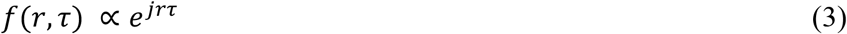

The identification of *f*(*r, τ*) above now enables us to find the spatial *UIR*(*r*), representing the spatial postsynaptic effects of an action potential effective over a fixed time epoch around the seizure onset ([−*T, T*]) as:

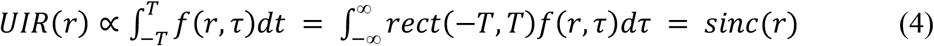

Interestingly, we find that both the temporal as well as the spatial ictal *UIR* are described by *sinc* functions, as shown by Equations (2) and (4).

### Temporal Cross-Correlation

If we represent the multi-unit spike train, with *N* spikes at times *t*_*i*_, as a series of delta functions 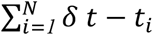, the temporal cross-correlation *C*(*τ*) between multi-unit spiking activity and the *LFP* is the spike-triggered average (STA):

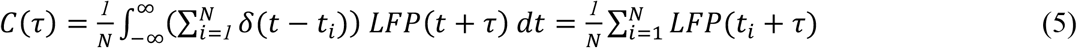

Thus, the average *LFP* in a window, determined by a positive or negative lag *τ*, around the spikes at *t*_*i*_. The temporal cross-correlations across interictal and ictal states are depicted in Fig. 2. In this figure, the black lines represent the cross-correlation signals, the red lines are the associated noise estimates, and the vertical dotted lines denote *t* = 0, i.e., the timing of the spike trigger. We build upon the data of both core and penumbral network states reported by Eissa et al. (2017) by extending the time base. The amplitude of the interictal signal is relatively weak (Fig. 2B, D) as compared to ictal state (Fig.2C, E). In the core, the interictal signal is dominated by a small triphasic wave (Fig. 2B). This signal transitions into a strong multiphasic signal during the ictal phase with a prominent negative phase (Fig. 2C). The *sinc*-like property of this ictal signal, can be appreciated at an increased time-scale (Fig. 3A).

**Figure 2:**
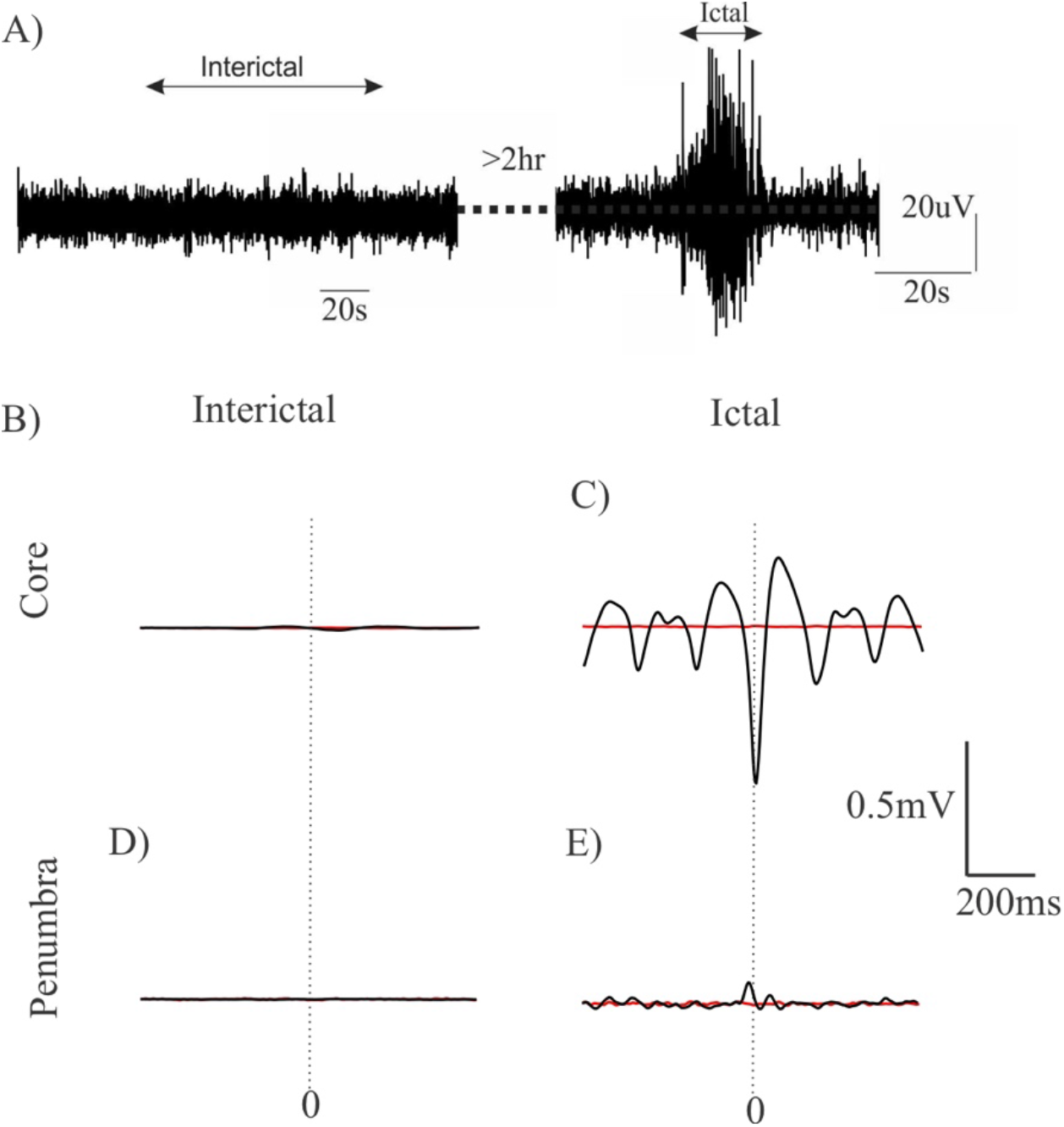
State dependent cross-correlations in different cortical territories. The black traces are the signals and the red ones represent the associated noise estimates. Vertical stippled lines represent the zero of the time-axis. A) Example signal trace of summed interictal and ictal LFP activity in the MEA. B – C) The cross-correlation signals in the core show an evolution towards a characteristic negative peak surrounded by oscillation during the ictal phase. As compared to the interictal cross-correlation, the amplitudes increase around the ictal phase. D – E) The cross-correlation data in the penumbra show ictal signals that are much smaller than the ones in the core. In contrast to the core, the small but dominant ictal peak polarity is positive.

**Figure 3:**
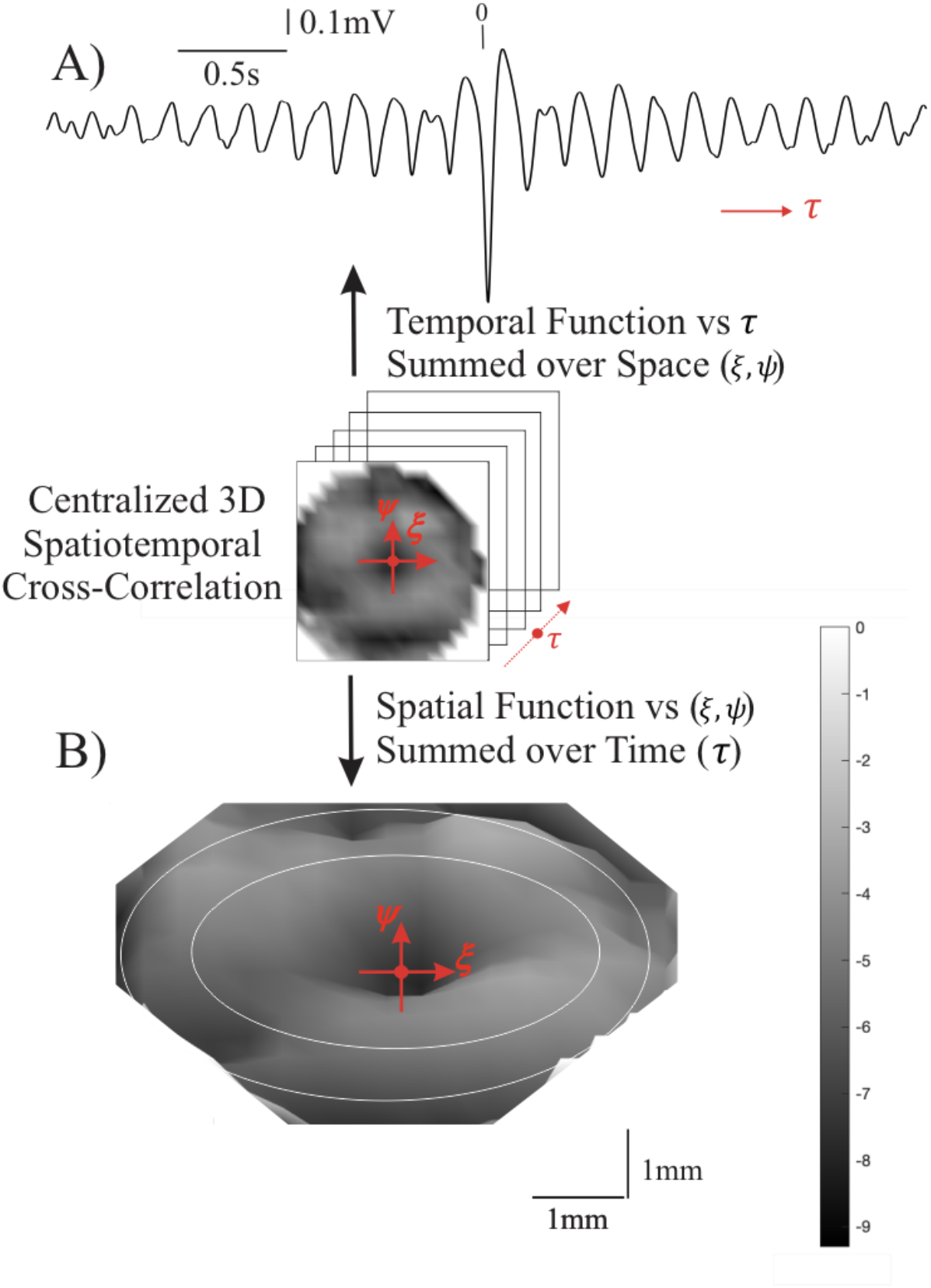
Properties of the ictal unit impulse response (*UIR*) function during a focal seizure. **A)** The temporal cross-correlation, *UIR*(*τ*), is calculated by summing the spatiotemporal cross-correlation over all spatial contributions (±3.6mm). **B)** A 3D view (azimuth = 0°, elevation = 70°) of the spatial *UIR*(*ξ, ψ*) summed over time *τ* = ±30ms. The center of the spatial *UIR* (*ξ, ψ* = 0,0) is indicated by the red dot. The two concentric circles are drawn to indicate that the center is surrounded by two rings. Note the (almost) radial symmetry of the *UIR*(*ξ, ψ*) function. For clarity, the corners are not depicted due to absence of sufficient measurement in that part of the area. Grayscale is in 0.1mV units.

In the penumbral network state, there are much smaller STA signals across all phases around the seizure (Fig. 2D, E). Here it should be noted that the cross-correlations of the penumbra in Fig. 2 are obtained by spikes evoked in that territory. In contrast, cross-correlations in the penumbra triggered by the spikes of the core are characterized by a significant oscillatory component, indicating the oscillatory activity in the penumbra is phase locked with spiking in the core and not with local spiking activity (3)(Fig. S3B).

### Spatiotemporal Cross-Correlation: Cartesian Coordinates

Next, we extended the commonly applied *temporal* cross-correlation between an action potential (spike) and the *LFP* by adding a *spatial* component. We used two spatial coordinates to represent the cortical surface. This produced a three-dimensional (two spatial dimensions and one temporal dimension) matrix of correlations values as a function of location (*ξ, ψ*) relative to spike position, and time (*τ*) relative to spike time (Methods, Fig. S1). A simplified analogy of this approach is the analysis of water ripples after throwing a stone into water. We can simulate multiple sources by dropping several stones from the same height, but across different times and locations in the horizontal plane above the water surface, resulting in a complex landscape on the surface of the water. To determine the contribution of a single stone to this landscape, we can take a field of view centered around individual stones. Averaging across all stones gives us the spatial pattern of activity associated with each stone. If we also include the time interval around each stone-drop, we get the stone’s characteristic spatiotemporal perturbation.

We obtained the temporal cross-correlation using the sum of the spatial values for each value of *t* and confirmed the result in (4) that the temporal cross-correlation resembles a *sinc* function. The central part (±2s) of the function reported in (4) is depicted in Fig. 3A. Similarly, we computed the cross-correlation as a function of location, (*ξ, ψ*), by summing over times *τ* = ±30ms (Fig. 3B). The spatial cross-correlation as depicted in Fig. 3B is determined by the extent and resolution of the size of the MEA and the computational procedure used (Methods, Fig. S1B). In the ictal phase, we observe a centrally located trough, surrounded by a pair of rings (indicated by the two white circles in Fig. S2B). The distance between the center and the region indicated by the inner circle is about 1.5 - 2mm and the distance between center and the region indicated by the outer circle is about 2.0 - 2.5mm. It can be seen in Fig. S2B that there is (almost) radial symmetry in the plane. The spatial cross-correlations during the interictal phase as well as the results obtained in the penumbra are smaller and show different patterns (Fig. S2).

### Spatiotemporal Cross-Correlation: Polar Coordinates

Under the assumption of radial symmetry in the cross-correlation, we converted the Cartesian coordinates (*ξ, ψ*) into polar coordinates (*r, θ*) and focused on the spatial relationship with respect to *r* (Methods, Fig. S1C). This enables us to depict the spatiotemporal properties in two dimensions, (*r, τ*) (Fig. 4A). A detail of that relationship is depicted in Fig. 4B and the summed values across this two-dimensional detail are plotted along the margins of the matrix in Fig. 4B. These summed values are the two cross-correlation components as a function of space and time (*r* and *τ*). Note that the bottom graph in Fig. 4B represents the central trough (*τ* = ± 30ms) of the function shown in Fig. 3A. As anticipated by the outcome in Equations (2) and (4), we observe a spatial component (left graph in Fig. 4B) that shows similarity to the pattern in the time domain, i.e., a central trough with smaller amplitude side lobes. Note that the limited resolution and range of the spatial component (*r* = ±3.6mm) is due to the size of the MEA and the computational procedure used (Methods, Fig. S1C).

**Figure 4:**
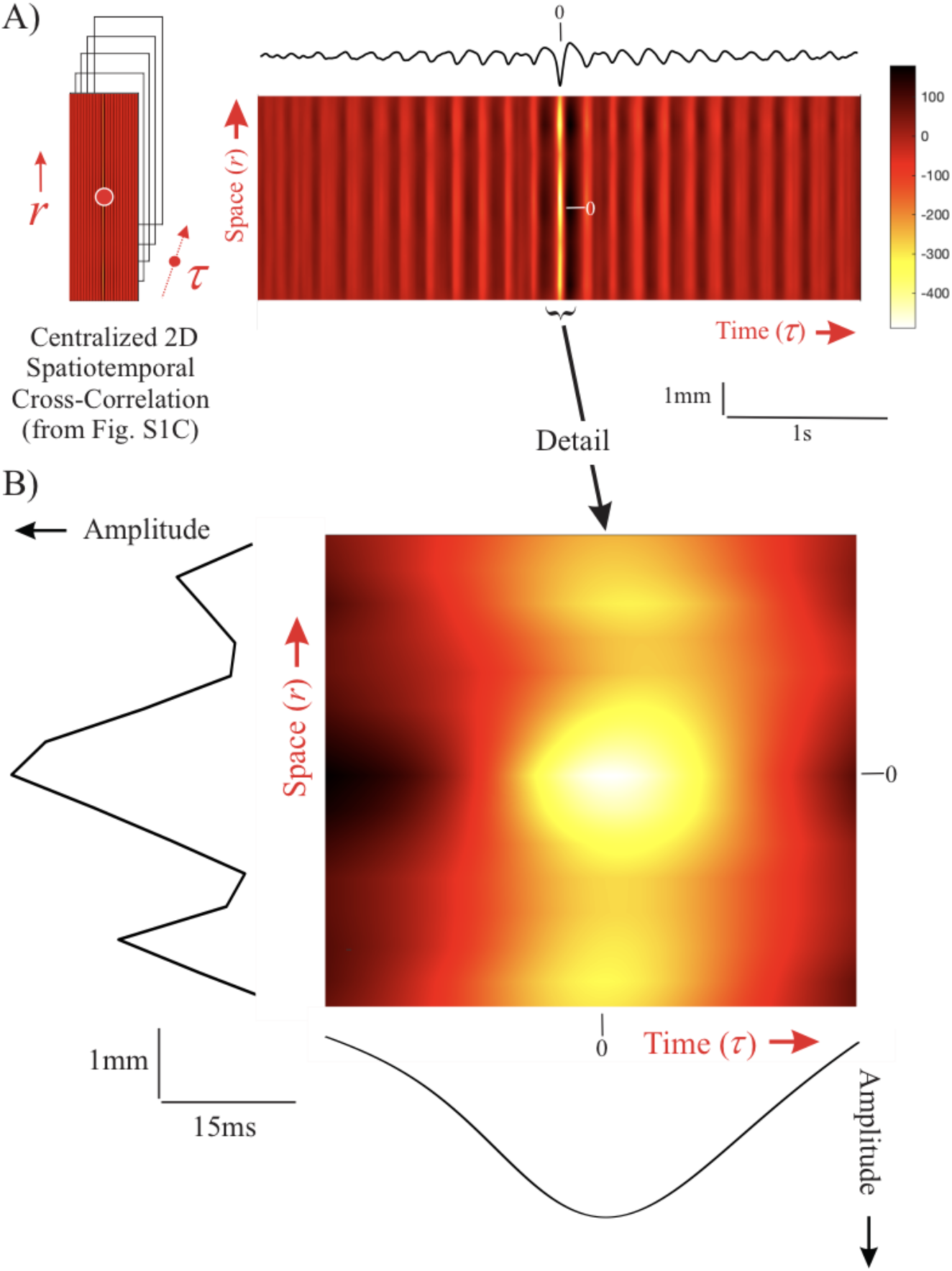
Centralized 2D spatiotemporal cross-correlation, *UIR*(*r,τ*). **A)** color representation of *UIR*(*r, τ*). The top trace, *UIR*(*τ*), is obtained by the sum of *UIR*(*r, τ*) over *r* (same as the signal in Fig. 2A). Amplitude and color scale are in arbitrary units.C **B)** Detail of the central part of panel A. The left margin shows the resulting wave from summation over time, generating *UIR*(*r*); the bottom margin depicts the resulting wave from summation over space, *UIR*(*τ*). The spatial *UIR* shows a distance between the peak and sidelobes of ∼2.0mm. Note that the bottom wave is (only) the central part of *UIR*(*τ*) shown in panel A.

## Discussion

The dynamics during human focal seizures (Movie S1) shows a complex relationship between action potential activity and *LFP*. As this may reflect a unique signature of seizures, with promise to further understanding of critical interactions relevant for their generation and propagation, we aim to determine their spatiotemporal characteristic across seizure states and cortical locations. Our model analysis (Equations (2) and (4), Fig. 1) and analyses from clinical recordings (Figs. 3, 4) lead us to conclude that during focal seizures the spatial cross-correlation can be approximated by a *sinc* function, similar to the previously reported time domain characteristic (4). In the following, we discuss the biological implications of our findings.

Under physiological conditions, an important contributor to the extracellular potential field is synaptic activity (6). Other contributors may include intrinsic membrane currents, gap junctions, neuron-glia interactions, and ephaptic effects (7, 8). While the relative contributions of these components during ictal conditions have not been fully elucidated, a non-zero cross-correlation between action potentials and *LFP*s is expected because synaptic currents are an important component of the compound activities during seizures. This is indeed shown by our spatiotemporal results (Figs. 2 - 4) and in the previously reported time-domain correlation (3, 4).

We adopt the interpretation of *LFP* polarity with respect to network state, as previously described in Fig. S10 in (3) and assign a net excitation to negative deflections and net inhibition to positive deflections. Accordingly, our temporal cross-correlation analysis (Fig. 2) shows that the ictal core’s action potential-LFP correlation at small lags is dominated by net excitation during the ictal phase.

When we apply the same polarity interpretation to the spatial domain (Fig. 3B), we observe that a ring of reduced excitation is present at a distance of ∼1mm around the excitatory center. The activity level in this excitatory center, representing the activity at the ictal wave, is excessively high, possibly due to saturation effects in the local inhibitory population (9). In turn, the ring of reduced excitation is surrounded by a second ring (at an additional distance of ∼1mm) where excitation is increased again (Fig. 3). This donut-shaped spatial cross-correlation is specific to the relationship in the core during the ictal phase (Fig. S2). This observation suggests that the ictal wave in the core territory, represented by the excitatory center (*ξ, ψ* = 0,0), creates an escape of (hyper)excitation via a jump across mm-range connectivity (Figs. 3 and 4). While this follows from the cross-correlation analysis, the question is whether there is a biological basis for this connectivity. First, in addition to short-range excitatory and inhibitory connections (within hundreds of μm), there are indeed excitatory mid-range connections (at the mm-scale) by axon collaterals within the grey matter in neocortex (Fig. 5A) (10–13). Second, we observed direct evidence that mm-range connections are invoked during the seizure. An example of this jump in action potential activity is depicted in the spatial plot in Fig. 5B (a snapshot of Movie S1): (I) indicates the location of the ictal wave front and (II) is an increase of neural firing over a gap of ∼2mm, which agrees well with the distance between the excitatory center and outer ring we observe in the donut shaped spatial cross-correlation depicted in Fig. 3. This pathological escape of uncontrolled excitation across cortex could be considered a candidate mechanism to recruit a critical mass of cortical neurons capable of sustaining the seizure.

**Figure 5:**
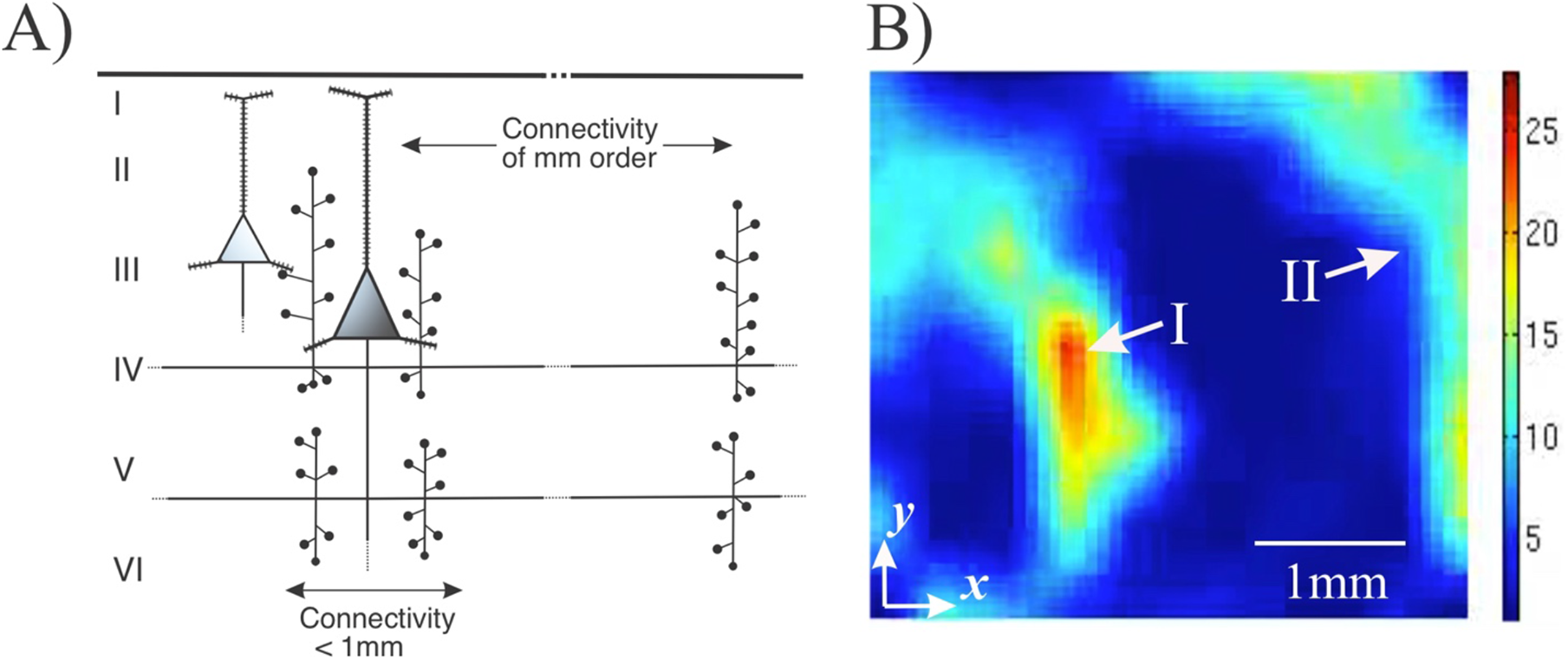
Mid-range excitatory neocortical connectivity by axon collaterals **A)** Diagram of grey-matter excitatory connections of a neocortical pyramidal cell showing the short-range connections (order of 100s of mm) and mid-range connections (order of mm) via the pyramidal cell axon collaterals (based on Nieuwenhuys, 1994, his Fig. 5). **B)** Snapshot of *Movie S1* depicting the propagation of ictal multi-unit action potentials across part of a Utah array. The area indicated by I is the propagating ictal wave, while the activation indicated by II shows activation at a mid-range distance of ∼2mm, indicating that the excitatory axon collateral connections are invoked for propagation of the ictal activity. Color scale: number of action potentials/100ms.

In sum, we examined and modeled spatiotemporal cross-correlations between network action potential activity and associated low frequency fluctuations of the local field potential in patients with focal seizures. We evaluated these correlation signals in two cortical territories (core and the surrounding penumbra) and across ictal and interictal states of the network. We show that: (1) the spatial and temporal components of the ictal network relate as a Fourier-transform-pair (Fig. 1); (2) the core and penumbra networks are functionally different in the interictal as well as ictal states (Figs. 2 and S2); (3) mid-range connectivity is involved in the outbreak of ictal hyper-excitation in the core area (Figs. 3, 4, and 5); (5) the correlations in core and penumbra show that the oscillations in the networks surrounding the core at cm-distance are not correlated by local spikes but by those in the core (Fig. S3). See SI for a summary of the mechanisms underpinning the focal seizure, using current findings combined with previous ones.

Our observations suggest that potential therapeutic strategies might target the spread of the wave’s excitatory effects via these collaterals. In this context, it is of interest that removal of horizontal interactions on a mm-scale has been a rationale for subpial transection in patients with intractable epilepsy (14).

## Materials and Methods

### Patients

Study participants (Table S1) at Columbia University Medical Center consisted of consented patients with pharmacoresistant focal epilepsy who underwent chronic intracranial EEG studies to help identify the epileptogenic zone for subsequent removal. Procedures were approved by the Internal Review Board committees at Columbia University Medical Center and The University of Chicago, Comer Children’s Hospital. The patients’ surgeries and treatment plans were not directed by or altered as a result of these studies.

### Signal Acquisition and Pre-Processing

A 96-channel, 4 × 4mm MEA (Utah array; Blackrock Microsystems) was implanted along with subdural electrodes (ECoG) with the goal of recording from seizure onset sites. Additional details of study enrollment and surgical procedures are described in (1). Signals from the MEA were acquired continuously at a sample rate of 30 kHz per channel (0.3-7500Hz bandpass, 16-bit precision, range ± 8 mV). The reference was epidural. Up to three seizures from each patient were selected for detailed analysis to avoid biasing the dataset from the patients from whom many seizures were recorded. Seizure recordings were categorized as core (seizure-generating area) or penumbra (surrounding unrecruited territory) using methods described in (1). Channels and time periods with excessive artifact or low signal-to-noise ratio were excluded.

Unit activity from the core was identified using filtered 0.3–3kHz signals with spikes defined as deflections ≥4 standard deviations below the mean. The low frequency component of the local field potential (*LFP*) activity across the array was created by averaging the artifact-free *LFP* activity from all micro-electrode signals filtered 2-50Hz. The averaged *LFP* procedure has been shown to generate signals that are representative of and comparable to nearby electrocorticography signals (3, 4).

### Cross-Correlation/Spike-Triggered Average and Signal Analysis

All signal processing and statistical analyses were performed in Matlab (Matlab, Natick, MA, USA). The evolution of the ictal neural activity in *Movie S1* shows the relationship between action potential activity and LFP.

The measured spatiotemporal relationship was determined as follows. First, as described in (3, 4), we detected the spikes in the multi-unit activity. Next, we collected the spatiotemporal data around each spike and translated the time and position of all associated LFPs relative to each spike’s time and position (Fig. S1A, B). Finally, we summed all translated data and computed the average at each time and position by dividing the sum by its number of contributions. Note that this position-dependent average is necessary because of the translation of the LFP’s axes, as not every position received the same number of contributions.

Evidence of radial symmetry of the spatial *UIR* (Fig. 2B) allowed conversion from (*ξ, ψ*) coordinates to polar coordinates (*r, θ*). By ignoring the minor deviations from radial symmetry, we focused on the spatial *UIR* relationship with respect to *r* (Fig. S1C), which enabled us to depict the spatiotemporal properties in 2D (Fig. 4). Furthermore, if we compute the sum across space, we obtain the temporal component of the *UIR*, while a summation over time *τ* generates the spatial component of the *UIR*. With these results, we can assess to what extent our model of the ictal network, a linear time-invariant (LTI) system with unit impulse response *UIR* ∝ *sinc*(*r, τ*), fits the data. A stepwise summary of the Methods can be found in the Supporting Information.

## Data Availability

All code used for statistical analysis and signal process is available upon request through the corresponding author. Data may also be requested through appropriate HIPAA-compliant channels and may be subject to institutional data sharing policies.

## Acknowledgments

We thank Drs. Stephan A. van Gils, Hil Meijer, Douglas R. Nordli Jr., and Jack D. Cowan for valuable discussion and suggestions. C.A.S., and W.v.D. were supported by NIH Grants R01 NS095368 and R01 NS084142. S.L and S.S.D. were supported by University of Chicago MSTP Training Grant T32GM007281.

## Supplementary Text

### 3D Cross-Correlation

The occurrence of action potentials across a seizing network is not experimentally controlled, unlike the scenario where the location and timing of the neuronal activities are evoked by external stimuli. The cross-correlation approach we used was designed to account for uncontrolled timing and location of spiking activity. In our analysis we assume propagation in the plane *x, y* as a function of time *t*. The spatiotemporal dynamics of the ongoing seizure can be characterized by the multi-unit action potential train (with *i* = 1, …, *N* action potentials), modeled by a series of unit impulses 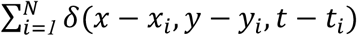, and the low frequency component of the local field potential (*LFP*). From Movie S1, we can conclude that the relationship between action potential activity and *LFP* seems a complex one, which motivated us to determine the spatiotemporal correlation *C*(*ξ, ψ, τ*) of the *LFP* with respect to the action potential:

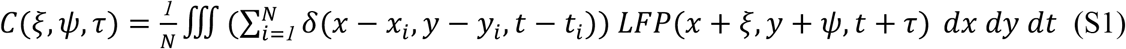

To evaluate this expression, we interchange the integration and summation operations and we integrate over the spatiotemporal domain of the ictal recording of the MEA:

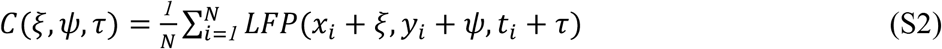

Note that here we add the spatial component to the commonly applied temporal cross-correlation/spike-triggered average, *C*(*τ*) (Equation (5)), with τ representing the lag to action potential timing. Our spatiotemporal result, *C*(*ξ, ψ, τ*), is the spatiotemporal cross-correlation/spike-triggered average with *ξ, ψ, τ* representing the space time coordinates relative to the position and timing of the action potential.

Using a similar approach as in (2) we now extend the model of the ictal network as a linear time invariant (LTI) system with the multi-unit action potential activity as input, the *LFP* as its output, and the network’s unit impulse response *UIR* defined as:

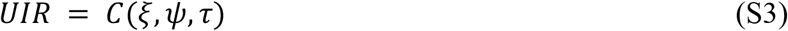

We now can recover the output *Z* of that LTI system using the convolution of the *UIR* and the network’s spikes:

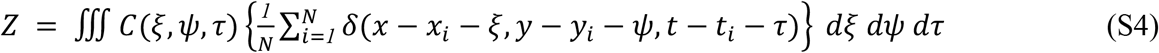

Note that we used the 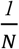 scaled version of the input here. Plugging in the expression for *C*(*ξ, ψ, τ*) results in:

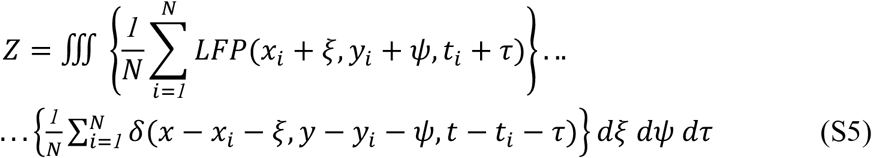

Exchange of the summation and integration operations and evaluation of the triple integral gives the model’s estimate of the spatiotemporal *LFP* from the LTI system:

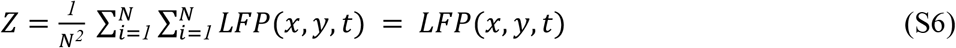

As shown in (2), the time domain component of this linear estimate produces a close approximation of the ongoing seizure activity with significant correlation (*p* < 0.01) between recorded and estimated activity.

### Signal Analysis

The measured spatiotemporal relationship was determined using the following steps (Fig. S1).

1. Each broadband signal of the 10×10 MEA was bandpass filtered for the low frequency component (2-50Hz) of the local field potential (*LFP*) and for spike detection (0.3–3kHz) (1, 2).
2. Spikes were detected in the multi-unit activity as negative deflections that exceeded four standard deviations of the filtered signal (1, 2).
3. For each spike the 10 × 10 frames of the *LFP* data were collected for ±*n* sample times representing ±5s around the spike time, and the timescale of the frames was set such that the spike occurred at time zero, *τ* = 0.
4. All *LFP* frames associated with a single spike were translated such that the spike location was at the origin of the new spatial coordinate system *ξ, ψ* = 0,0. Note that this spatial translation is necessarily spike specific because spikes do occur at different locations.
5. Next, the translated 10 × 10 × (2*n* + 1) frames were put into a three dimensional 19 × 19 × (2*n* + 1) configuration with the spatiotemporal origin (*ξ, ψ, τ* = 0,0,0) is at position 10,10, *n* + 1. This step was done to keep the *LFP* frames compatible across spikes.
6. For each spike, these frames were summed into a three dimensional 19 × 19 × (2*n* + 1) matrix.
7. For each position in the 19 × 19 × (2*n* + 1) matrix, the total number of contributions *N* was counted.
8. Finally, to obtain the spatiotemporal cross-correlation, the sum obtained in step 6 was divided by the *N* obtained in step 7 for each position. This resulted in the discrete spatiotemporal estimate of *C*(*ξ, ψ, τ*), as shown in Equation (S3).

To evaluate the signal-to-noise ratio (*SNR*) of the averaged results, we estimated the residual noise using the plus-minus averaging approach. We implemented this by employing the above eight steps while keeping two three-dimensional 19 × 19 matrices: one summed the even contributions for each location and the other summed the odd ones. To obtain the averages for the odd and even components, each position in the matrix was then divided by its number of contributions. The sum of the even and odd averages is the same result obtained in step 8 above. In contrast, the difference between the even and odd averages cancels the consistent component (i.e., the signal) while preserving the random noise estimate (3, 4). The *SNR* was estimated by computing the root mean square (*rms*) of the signals and the *rms* of their noise estimates, leading to a signal-to-noise ratio,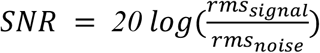. Average ratios for the cross-correlations across space and time all were >30dB.

### Mechanisms involved in focal seizures

By combining current and previous findings on ictal dynamics, we can outline the following summary for an evolving neocortical focal seizure. At the micro and meso-scales, an ictal wave of action potential activity propagates at a velocity of ∼1mm/s by invoking excitation via the local connections over distances < 1mm. This wave of hyperexcitation propagates locally when the inhibition in front of this wave fails to constrain the excitation (1, 5, 6). In this context, it is interesting to note that this propagation process is compatible with the evolution of the clinically observed Jacksonian march first described by Hughlings Jackson in 1870 (e.g. 7). We now find evidence that, in addition to the slow propagation process, the ictal wave excites cortical areas farther than 1mm away, probably via axon collaterals within the grey matter, which allows excitation to ‘escape,’ and enables recruitment of additional cortical territory. This activation of areas > 1mm away might also explain modular propagation of ictal activity, a property previously observed in experimental seizures (8). At the macro-scale, white matter intracortical connections are invoked, spreading ictal activity across a cm-sized territory. The activity in this macroscale territory is still highly correlated with the action potential activity in the ictal wave located in the core rather than the local action potential activity located in the penumbra (Fig. S3) (1). In addition, while local inhibition fails at the ictal wave front, longer range inhibition remains intact and plays a critical role in sustaining the synchronous oscillatory component of the ongoing seizure at the macroscale (1, 2).

## Supplementary Tables and Figures

**Table S1.** Patient demographics (1)

|  | **Patient (age/gender)** |  |  |  |
| --- | --- | --- | --- | --- |
| **Patient features** | **Patient 1 (25 y old/female)** | **Patient 2 (19 y old/female)** | **Patient 3 (30 y old/male)** | **Patient 4 (39 y old/male)** |
| Implant location | Left lateral and subtemporal | Right lateral and subtemporal, parietal, occipital | Left lateral frontal, mesial frontal, temporal | Left lateral and mesial frontal |
| MEA location | Left inferior temporal gyrus 2.5 cm from anterior temporal pole | Right posterior temporal, 1 cm inferior to angular gyrus | Left supplementary motor area, 3 cm superior to Broca's area | Left lateral frontal 2 cm superior to Broca's area |
| Seizure onset zone | Left basal/anterior temporal (including MEA site) | Right posterior lateral temporal, (including MEA site) | Left supplementary motor area (including MEA site) | Left frontal operculum (3 × 3-cm cortical area, including MEA site) |
| Days recorded | 5 | 28 | 4 | 4 |
| No. of seizures analyzed | 3 | 1 | 3 | 3 |
| Seizure type(s) | Complex partial | Complex partial with secondary generalization | Complex partial/tonic | Complex partial |
| Pathology | Mild CA1 neuronal loss; lateral temporal nonspecific | Nonspecific | N/A (multiple subpial transections performed) | Nonspecific |

- N/A, not applicable.

**Figure S1:**
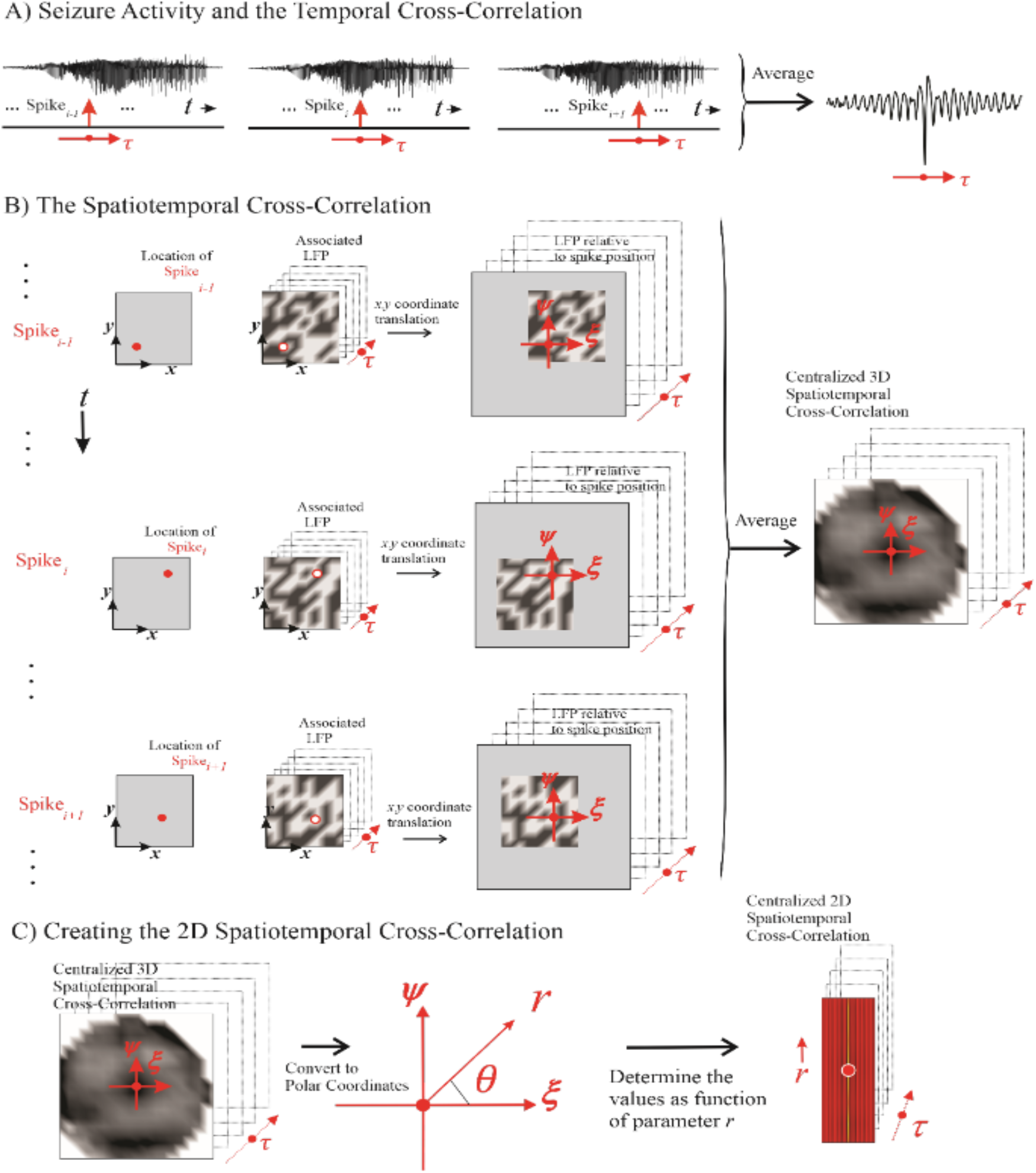
Mesoscale spatiotemporal cross-correlation between spiking activity and the low frequency component of the local field potential (*LFP*) during a human focal seizure. **A)** During seizure activity, the *LFP*s within the electrode array’s area (the summed *LFP* the MEA is depicted in the upper trace) are associated with a multi-unit action potential train. For clarity, only single action potentials, i.e., spikes (red impulse), out of this train are depicted in the lower traces. The *LFP*’s relationship to the spike is considered over time *τ* relative to the spike events. **B)** For each spike (left column, ‘Location of Spike…’, red dot), its associated spatiotemporal *LFP* (middle column, ‘Associated *LFP*’, grey scale) is determined. The red circle in the ‘Associated *LFP*’ column indicates the spike position on the *LFP* map. Next, the *x, y* axes of the *LFP* are translated into the *ξ, ψ* axes, such that the associated spike position (red circle) is at the origin (right column, ‘*LFP* relative to spike position’). Finally, the results in the right column are averaged (‘Centralized 3D Spatiotemporal Cross-Correlation’), a matrix that contains the centralized spatiotemporal cross-correlation (i.e., spike-*LFP* correlation). Note that all axes (*ξ, ψ, τ*) are translated to reflect the *LFP* relative to both spike position and timing. Also note that due to the translation procedure for each action potential, each position in the centralized spatiotemporal cross-correlation may have a different number of contributions – therefore each position is averaged individually. Due to the translation procedure, the corners of the average are undefined (indicated by the white areas at the corners). **C)** The Cartesian coordinates (*ξ, ψ*) of the end result in panel B are converted into polar coordinates (*r, θ*). Focusing on the radial component we convert the 3D into a 2D matrix (‘Centralized 2D Spatiotemporal Cross-Correlation’).

**Figure S2:**
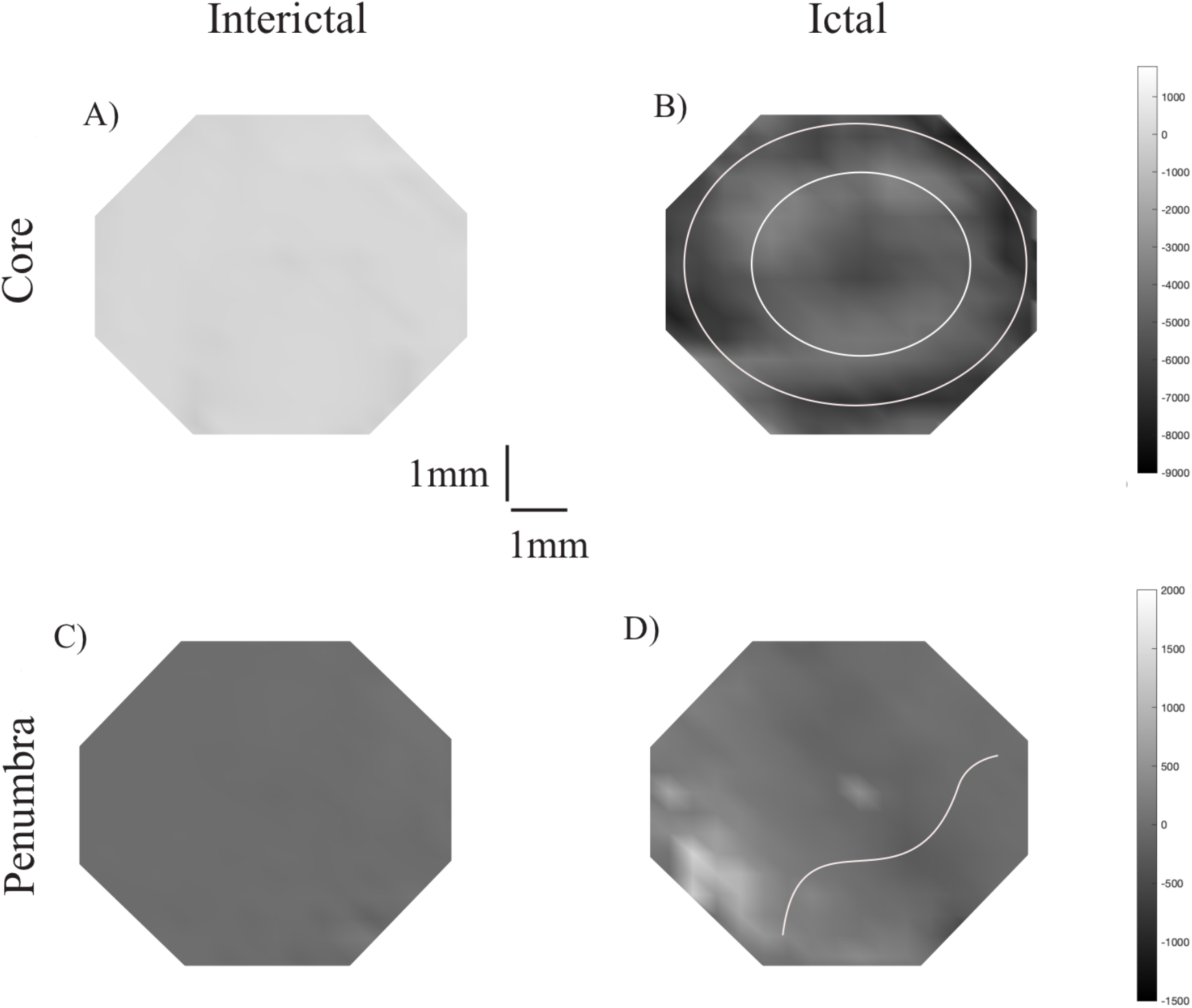
Spatial cross-correlation in core and penumbra territories. The spatial cross-correlation in core (A-B) and penumbra (C-D). These signals represent the same spike-LFP relationship as depicted in Figure 2B. Grey scale is in 0.1μV units. In Panel B it can be seen that the donut shaped ictal signal in the core is strong as compared to interictal phase. The two rings surrounding the center are indicated by the white circles in panel B). The penumbra dynamics is markedly different and are also much smaller in amplitude (note the different scales between core and penumbra panels). The ictal phase in the penumbra suggests a wave-like distribution of the landscape (indicated by the white wave in panel D).

**Figure S3:**
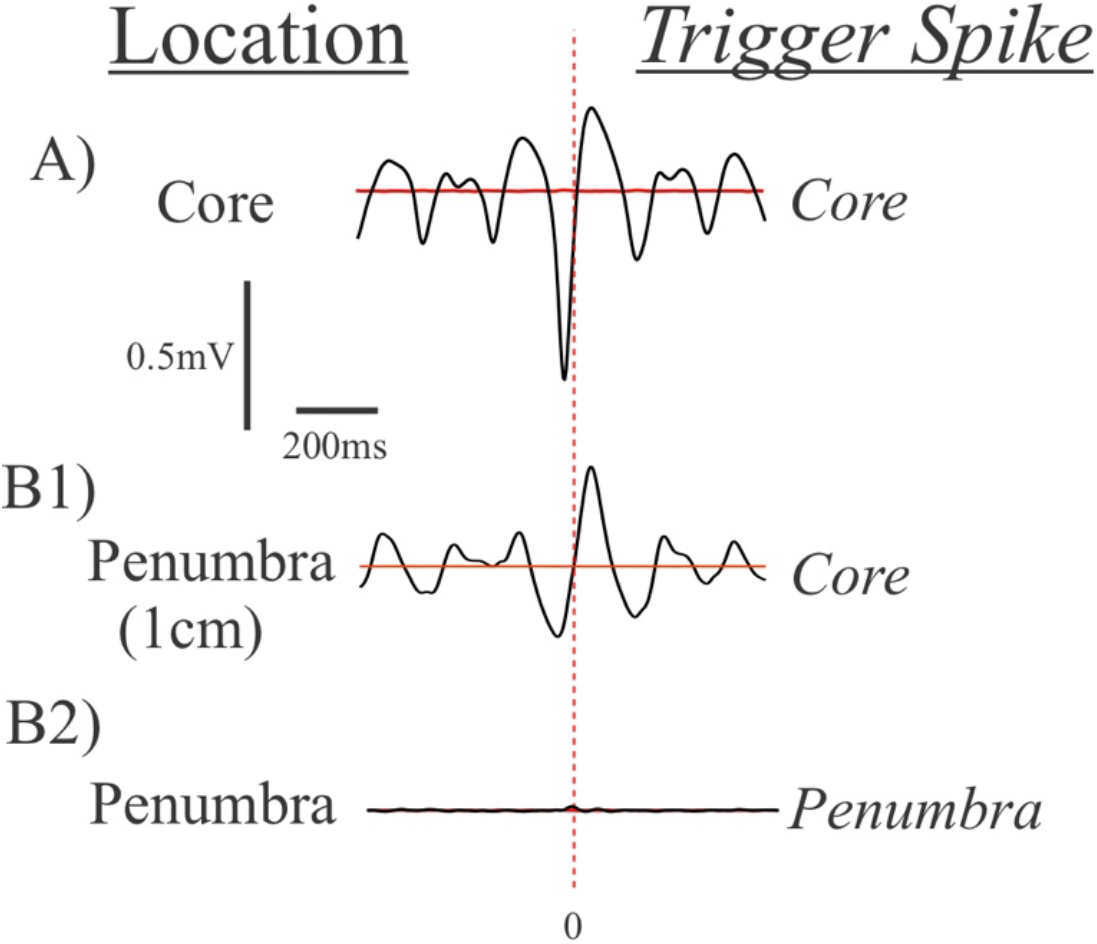
Spike-triggered averages (temporal cross-correlations) in core and penumbra territories. **A)** The spike-triggered average in the core. **B)** The spike-triggered average in the penumbra. B1) is triggered by spikes in the core at a distance of 1cm, while B3) shows a spike-triggered average using the local spikes in the penumbra.

**Figure S4:**
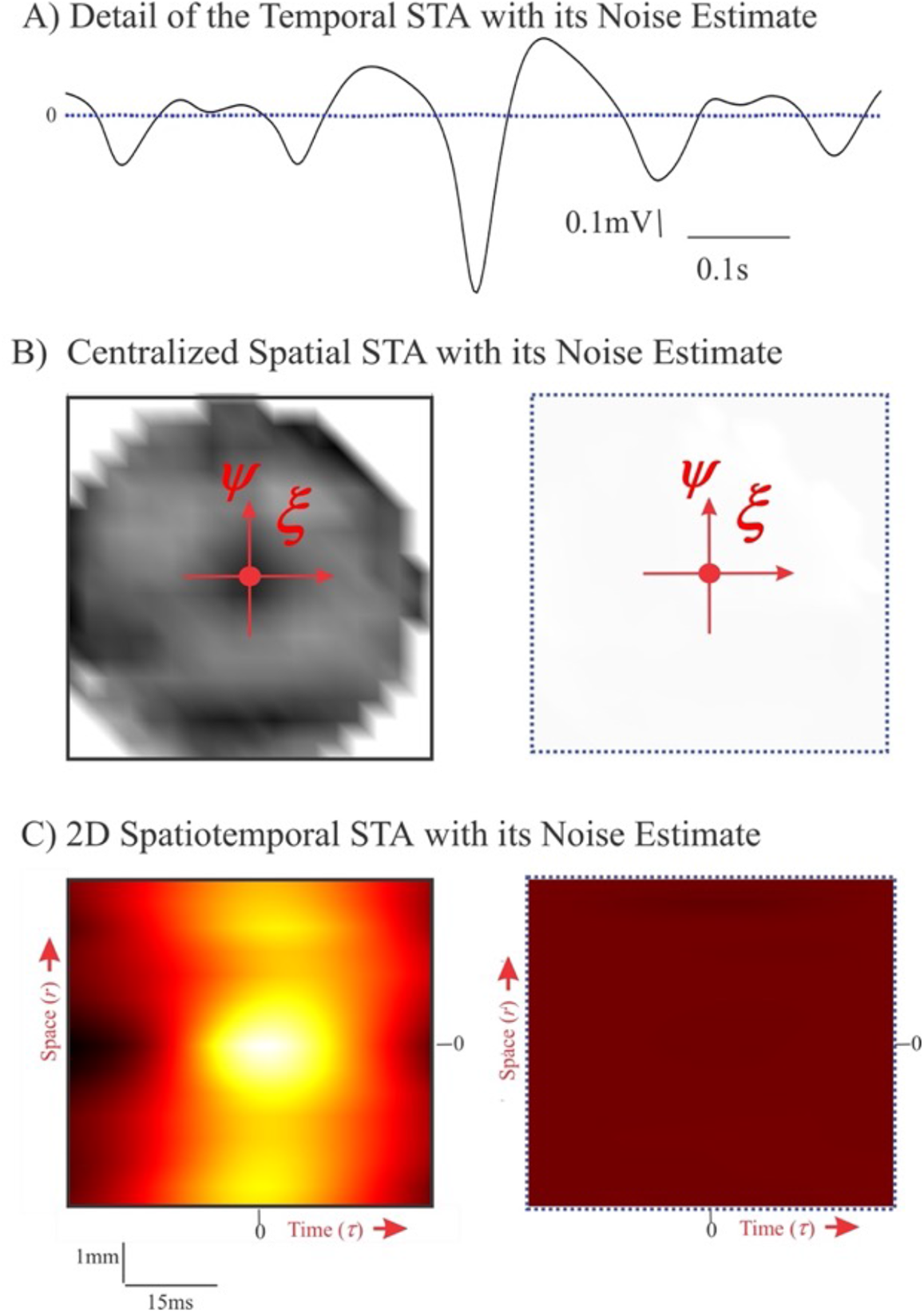
Noise estimates of the spatiotemporal cross-correlation of the data in Figs.3 and 4. **A)** Detail of the spike-triggered average (temporal cross-correlation) from Fig. 3A (solid, black) and its noise estimate (dashed, blue). The signal-to-noise ratio of the depicted data is 45dB. **B)** The spatial cross-correlation depicted in Fig.3B (left) and its estimated noise component (right). The location specific signal-to-noise ratios of the depicted data range is 26 – 80dB, with an average of 38dB. The units for the grey scale are identical for both maps and identical to the scale in Fig. 3. **C)** The 2D version of the spatiotemporal cross-correlation from Fig. 4B (left) and its noise estimate (right). The signal-to-noise ratio of the depicted data is 39dB. The units for the color scale are identical for both maps and the same as in Fig. 4.

## Legend for Movie S1

Movie of the multi-unit activity depicting the propagation of the ictal wavefront (left panel) and the associated 2-50 Hz low frequency component of the *LFP* (right panel) from the MEA from Patient 2 are shown. The top trace shows *LFP* activity from a single representative MEA channel. Five seconds beginning 0.5s before onset are shown. With permission from (5).

